# Adherence to Healthy Lifestyle and Associated Factors among Hypertension Patients Under Follow-up at Public Hospitals in Northwest Amhara, Ethiopia: A multi-center cross-sectional study

**DOI:** 10.1101/2025.06.30.25330583

**Authors:** Melkamu Zeleke, Getalem Aychew Beyene, Telake Azale, Alemakef Wagnew, Minichil Genet, Hibret Getaneh Zeleke, Dagmawi Zemene, Tinbite Esayas Desta, Achenef Asmamaw Muche

## Abstract

**Objective:** *The aim of this study was to assess adherence to a healthy lifestyle and associated factors among hypertension patients at selected public hospitals in Northwest Amhara, Ethiopia*.

**Study design:** *A multi-center hospital-based cross-sectional study design was conducted*.

**Study setting:** *The study was conducted at primary and general hospitals in Northwest Ethiopia*.

**Primary and secondary outcome measures:** *Adherence to a healthy lifestyle was assessed with respect to adherence to diet, physical exercise, smoking, and moderation of alcohol consumption, specifically assessed with the 4-item Fast Alcohol Screening Test as the primary outcomes, and factors associated with adherence to a healthy lifestyle were secondary outcomes*.

**Participants:** *Eight hundred and forty adult hypertension patients were involved in this study; of these, four hundred forty-eight were men and three hundred ninety-two were women. Systematic sampling techniques were used to select study participants. Data were collected using structured interviewer-administered questionnaire and chart review. We analyzed the data using SPSS V.20 statistical software. An adjusted odds ratio with a 95% confidence interval and a p-value <0.05 was used to determine predictors of adherence to a healthy lifestyle*.

**Result:** *The overall good adherence in this study was 21.7% (95% CI: 19.0 to 24.6). Being female (AOR = 2.03; 95% CI: 1.38 to 2.98), having primary education (AOR = 2.07; 95% CI: 1.06 to 4.03), a duration of hypertension diagnosis of 2-4 years (AOR = 3.14; 95% CI: 1.83 to 5.39), having good knowledge (AOR = 3.06; 95% CI: 2.02 to 4.64), and getting good social support (AOR = 2.89; 95% CI: 1.97 to 4.25) were significant predictors of good adherence to healthy lifestyle modification practices*.

**Conclusion:** *The overall good adherence to recommended healthy lifestyle modification practices was low. Good adherence was associated with being female, having primary education, having been diagnosed with hypertension for a period of time, having good knowledge, and receiving social support. **Keywords:** hypertension; healthy lifestyle; adherence; Ethiopia*.

**STRENGTH AND LIMITATION OF THE STUDY:**

❖ *Eleven items on the Likert scale and the Duke Social Support and Stress scale were used to assess level of social support, which was a globally accepted protocol*.
❖ *The 4-item Fast Alcohol Screening Test (FAST), which is the short version of the alcohol disorders identification test (AUDIT), was also used to assess adherence to alcohol consumption*.
❖ *It is challenging to determine the causal linkage between adherence to a recommended healthy lifestyle and associated factors due to the cross-sectional nature of the study design*.
❖ *Regarding diet intake, it was challenging to quantify exactly what they consumed because the diet measuring questionnaire was subjective*.
❖ *The other limitation of the study is that the questionnaire employed in this study might be prone to social desirability and recall bias*.

## INTRODUCTION

Hypertension (HTN) is defined as a persistent increase in systolic blood pressure (BP) ≥140 and/or diastolic BP ≥90 mmHg in adults aged 18 years and older, according to the World Health Organization (WHO).^1^ High blood pressure is a serious global public health concern due to its high incidence, significant increase in death, and correlation with both cardiovascular disease and chronic renal illness.^1-5^ Globally, high blood pressure is the leading preventable cause of mortality and disability in the world, affecting 1.39 billion adults worldwide. Of these, 349 million people live in high-income countries, while 1.04 billion live in low- and middle-income countries.^6^ Even though it is a worldwide public health concern, more than three-fourths of deaths occur in populations in low- and middle-income countries, which account for nearly 80% of deaths in countries with inadequate health systems.^6,7^

In addition, hypertension accounts for 7.6 million fatalities worldwide each year.^8^ The highest prevalence of high blood pressure was found in the African region, where it was 46% in both sexes, while the lowest prevalence was found in the WHO region of the Americas, where it was 35% in both sexes.^9^ In Ethiopia, based on a systematic review and meta-analysis found in 2020, the prevalence of hypertension was found to be 27.9%. The other studies also indicated that the prevalence of hypertension was 27.9%, 25.35%, and 15.36% in Dabat District, Gondar Town, Addis Ababa, and Tigray Region, respectively.^10,11^ In 2013, Ethiopia launched national non-communicable disease (NCD) program, which included hypertension, and the hypertension prevention and control strategic plan and guidelines were also developed in 2014 and 2016, respectively. The national guideline recommended both pharmacological and non-pharmacological treatments for hypertension patients.^12^ Healthy weight, physical exercise, salt limitation, smoking cessation, high fruit and vegetable intake, low-fat and oily diets, and limiting alcohol consumption are all recommended healthy lifestyle practices by the guideline.^13^ One of the most important aspects of hypertension therapy is maintaining a healthy lifestyle. This includes the patient’s ability to adopt a diet, frequent exercise, alcohol moderation, and quitting smoking.^14-15^

An effective lifestyle modification can reduce the BP as much as a single antihypertensive medication. Adherence to lifestyle modification is still poor, but adherence to anti-hypertensive medication is relatively high, at about 67%.^16^ Despite the evidence for the importance of a healthy lifestyle, studies in Ethiopia showed that hypertension patients’ adherence to healthy lifestyle recommendations is poor.^16-18^ About 70% of people with hypertension who simply get pharmaceutical treatment have uncontrollable blood pressure. ^19^ In Ethiopia, it is estimated that 48% of hypertension patients have uncontrolled blood pressure (BP).^20^ A study on the adherence to lifestyle modification practices carried out in the Amhara region of Dessie and Bahir Dar found that the respective rates of adherence were only 23.6% and 18.32 respectively.^18,21^

Epidemiological data on the magnitude of adherence to a healthy lifestyle is important for planning and implementing non-pharmacologic hypertension management. Apart from existing interventions to address the problem by the government of Ethiopia and other actors, limited research was done on the magnitude and associated factors of adherence to a healthy lifestyle practice by hypertension patients in Ethiopia, particularly in a study area. Thus, this study aimed to determine adherence to healthy lifestyle and its effect to control hypertension among hypertension patients at selected public hospitals in Northwest Amhara, Ethiopia.

## MATERIAL AND METHODS

### Study design, setting and period

The objective of the study was *to assess adherence to a healthy lifestyle and associated factors among hypertension patients at selected public hospitals in Northwest Amhara, Ethiopia.* A hospital-based cross-sectional study was conducted from August 20 to September 25, 2022, in selected public hospitals across four zones in Northwest Amhara: Central Gondar, North Gondar, South Gondar, and West Gondar. All four zones included in this study contain a total of 41 districts. As of the 2007 national census prediction, the study area had a total population of 6, 219, 763. In the respective zones, there are a total of 22 primary hospitals, one specialized teaching referral hospital, one referral hospital, and two general hospitals that provide chronic follow-up care for chronic disease patients.

The current study was conducted in two general hospitals and two primary hospitals, namely Debark General Hospital (DGH), Metema General Hospital (MGH), Wogera Primary Hospital (WPH), and Dembiya Primary Hospital (DPH) in North Gondar, West Gondar, and Central Gondar Zones respectively.

### Study population

The source population was all adult hypertensive patients receiving follow-up care at public hospitals in the four zones of Northwest Amhara. The study populations were all adult hypertensive patients who were on follow-up care at the selected hospitals during the study period.

### Ethics approval

The University of Gondar’s Institutional Review Board (IRB) granted ethical approval. A support letter was obtained from the University of Gondar, College of Medicine and Health Science, Institute of Public Health, and submitted to the respective hospitals.

### Patient and public involvement

Patients and/or the public were not involved in the design, or conduct, or reporting, or dissemination plans of this research.

### Eligibility criteria

All hypertensive patients aged o ≥18 who had been on follow-up care at the selected public hospitals for at least 6 months and visited chronic follow-up department for follow-up visits while receiving antihypertensive treatment during the study period. Patients who were seriously ill at the time of data collection and unable to respond to the questions, patients with cognitive impairment, patients with hypertensive urgency or emergency, patients who declined to participate in the study, and incomplete medical records or missing demographic or blood pressure data were all excluded.

### Sample size determination and sampling procedure

The sample size for the first objective was calculated using a single population proportion formula, with the following assumptions: a 95% confidence level (CI), 46.4% of the adherence rate to lifestyle modifications among hypertensive patients in Yekatit 12 Hospital in Addis Ababa, a 5% margin of error (d), a design effect of 2, and a 10% contingency for the non-response rate. Then, the final sample size becomes 840. ^22^

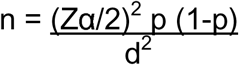

Where: n = the required sample size, p = proportion of adherence to lifestyle modification, Z α/2 = the standard normal distribution curve value for a 95% confidence interval, which is equal to 1.96, d = margin of error (5%), and α = probability to reject the true null hypothesis.

The sample size for the second objective is calculated using the double population proportion formula and some determinants for adherence to a healthy lifestyle, based on the following assumptions:

**P1**=% outcome in unexposed group

**P2**= % of outcome in exposed group

**OR**=Odds ratio

**CI**=Confidence Interval

The largest sample size from the second objective was 348, taking OR =0.53, power= 80, zα/2 95%CI= 1.96, ratio= 1, p1=53.3 and p2= 37.8. ^23,24^ By taking the larger number, the final sample size for this study was 840.

In the Northwest Zones of the Amhara region, there are 26 public hospitals that provide chronic follow-up care for chronic disease patients. A simple random sampling technique was employed to select four public hospitals. The research was conducted in public hospitals with chronic follow-up clinics. The sample was selected using stratified sampling techniques by taking each hospital as a stratum, and in each stratum, the number of study units was allocated proportionally based on the number of patients on monthly follow-up. The study participants were selected using systematic sampling techniques and proportionally allocated according to population size after determining the sampling interval value (N/n), where N is the total population during the study period in the study area and n is the sample size determined for the specific area. The first participants were selected using the lottery method, and data was collected until the required sample size was achieved according to the monthly follow-up of patients in each hospital’s proportional allocation (Fig. 1). ^25^

**Figure 1.**
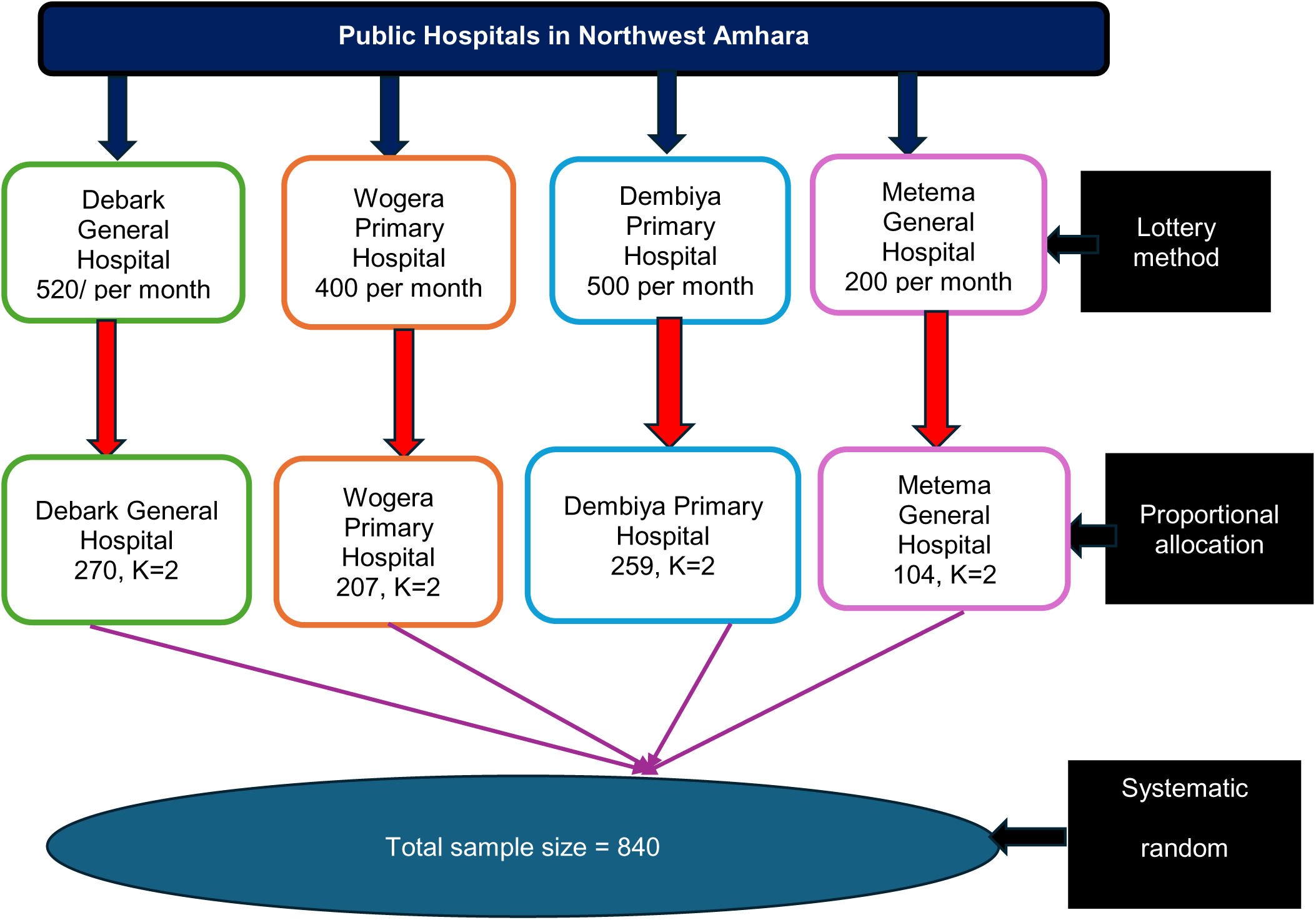
Schematic presentation of the sampling technique used to select the study participants from public hospitals of Northwest Amhara, Ethiopia.

### Study variables Dependent variable

Adherence to healthy lifestyle: - adherence to diet, physical exercise, smoking, and moderation of alcohol-consumption related recommendations.

### Independent variables

**Socio-demographic variables:** age, sex, marital status, place of residence, religion, educational level, occupational status and monthly income

**Clinical factors:** presence of comorbidities, types of anti-hypertensive medication intake, family history of hypertension (HTN), body mass index (BMI), and frequency of follow-up

**Individual factor:** knowledge about hypertension

**Social support:** support from families and non-family members of the society

### Operational definitions

**Adherence to healthy lifestyles:** Respondents who were adherent to the diet, physical exercise, smoking, and moderation of alcohol consumption-related recommendations. In this study, the respondents who adhered to all four healthy lifestyle recommendations were considered good adherents and poor adherents if adherence to any one or more components was missing.^22^

**Diet-related adherence:** those who report that they usually or always consume a diet rich in fruits, vegetables, and whole grains and never or rarely consume salt, sugar, or saturated fat were taken as adherent or otherwise non-adherence to the diet.^18^

**Physical exercise-related adherence:** Respondents that performed moderate physical activity exercise for 30 minutes (e.g., brisk walking, jogging, cycling) for at least 5 days a week were considered adherent or otherwise non-adherent to physical exercise.^13^

**Alcohol-related adherence:** Respondents who report that they either never consumed alcohol or whose total score on the 4-item Fast Alcohol Screening Test (FAST), which is the short version of the alcohol disorders identification test (AUDIT), is <3, were taken as adherent or otherwise non-adherent to moderation of alcohol consumption.^16^

**Smoking-related adherence:** Participants who report that they either never smoked or stopped smoking before 30 days were taken as adherent or otherwise non-adherent to smoking.^26^

**Social support:** support received from family and non-family members. Respondents whose score was above the mean value on the 11 items on the Likert scale and the Duke Social Support and Stress scale were taken as having good social support.^18^

**Comorbidities:** hypertension patients with one or more medical conditions in addition to HTN.

**Knowledge about HTN:** Respondents with scores above the mean value on the 8-item modified hypertension evaluation of lifestyle and management (HELM) scale were taken as having good knowledge or otherwise poor knowledge about hypertension.^18^

### Data collection tool and procedures

Data were collected using a structured interview-administered questionnaire and chart review. The questionnaires were adapted from validated scales and previous similar studies and the ministry of health NCD guidelines of Ethiopia and modified for this study.^10,16,18,22^ Data abstraction format was designed and prepared to complete patients’ information from their record sheet. The questionnaires were first prepared in English, then translated to the local language, Amharic, by multi-lingual experts, and then translated back to English to maintain its consistency. ^27^ Each designated data collector received the patient files from the assigned health care professional based on daily patient follow-up at respective hospitals. Then necessary data were completed using the developed data abstraction format. Patients were interviewed to obtain socio-demographic, knowledge about hypertension, disease related, level of social support, and lifestyle related information (questionnaires attached as supplementary file 1). The participants BP was measured using a mercury sphygmomanometer with adult cuff size and stethoscope after the patients’ rested for 10 minutes and was measured two times in one visit, if the result has difference take the average of the two measurements. The weight in kilograms and height in centimeters of participants were measured barefoot and wearing light clothing with a standing weight and height scale; the body mass index (BMI) is calculated as weight in kilograms divided by height in meters squared. Four trained nurses collected data from different hospitals while being supervised by two public health specialists with expertise in supervising research pertaining to hypertension.

### Data quality control

To ensure the quality and clarity of the questionnaire and the data abstraction format, a pre-test was conducted among 5% (42) of the entire sample in a chronic hypertension follow-up clinic at Adis-Zemen Primary Hospital prior to the actual data collection. In addition, two days of training were given for data collectors and supervisors on the overall data collection procedures and the techniques of interviewing, including how to extract patients’ information from the patient’s record sheet. A reliability test (Cronbach alpha) was performed to check the internal consistency for the Fast Alcohol Screening Test (FAST) which was 0.798; the Duke Social Support and Stress scale, which was 0.778, and knowledge about hypertension, which was 0.76. The collected data were checked for legibility of handwriting, completeness, consistency, accuracy, and clarity by the principal investigator and supervisors on a daily basis.

### Data processing and analysis

Epi Info Version 4.6 was used to enter, clean, and export to Stata Version 14 for analysis. To verify correctness, consistency, and the lack of missing values, the data was cleaned using frequency and cross-tabulation. Furthermore, the chi-square assumption was checked before the logistic regression analysis. Binary logistic regression analysis was used to determine the association between dependent and independent variables. The statistical assumptions for binary logistic regression were assessed, and no significant violations were detected.

The Hosmer-Lemeshow test was checked for model goodness of fit (p = 0.26) and the variance inflation factor (VIF) was done to check for multicollinearity. During the multivariable logistic regression analysis, the potential effects of confounders were controlled. Variables with a p-value < 0.2 during bivariable analysis were fitted to multivariable analysis. An adjusted odds ratio with a 95% confidence interval and a p-value <0.05 was used to determine predictors of adherence to a healthy lifestyle.

## RESULTS

### Socio-demographic characteristics

A total of 840 adult hypertensive patients were included in this study, with a response rate of 100%. The median age of the respondents was 50 years, with a ±16 interquartile range. More than half, 448 (53.3%) of the respondents were male, and the majority, 601 (71.6%) were married. Nearly half of the respondents, 436 (52%), were orthodox by religion. Out of the respondents, about one-third, 287 (34.2%) were unable to read and write, and about 399 (47.5%) were employed (Table 1).

**Table 1:** Socio-demographic characteristics of adult hypertension patients under follow-up at the selected public hospitals in Northwest Amhara, Ethiopia, 2022 (n=840)

| Variables |  | Frequency(n=840) | Percent (%) |
| --- | --- | --- | --- |
| Sex | Female | 392 | 46.7 |
|  | Male | 448 | 53.3 |
| Age | 22-35 | 106 | 12.6 |
|  | 36-45 | 208 | 24.8 |
|  | 46-55 | 265 | 31.6 |
|  | >55 | 261 | 31 |
| **Marital status** | Married | 601 | 71.6 |
|  | Single | 63 | 7.5 |
|  | Divorced | 69 | 8.2 |
|  | Widowed | 107 | 12.7 |
| **Residency** | Urban | 439 | 52.3 |
|  | Rural | 401 | 47.7 |
| **Religion** | Orthodox | 436 | 52 |
|  | Muslim | 280 | 33.3 |
|  | Protestant | 81 | 9.6 |
|  | Catholic | 43 | 5.1 |
| **Educational level** | Unable to read and write | 287 | 34.2 |
|  | Able to read and write | 149 | 17.7 |
|  | Primary | 109 | 13 |
|  | Secondary | 77 | 9.2 |
|  | College or university | 218 | 25.9 |
| **Occupational status** | Unemployed | 441 | 52.5 |
|  | Employed | 399 | 47.5 |
| **Monthly income** | <3000 | 243 | 28.9 |
|  | ≥ 3000 | 597 | 71.1 |

### Clinical characteristics of the participants

Of the respondents, about 243 (28.9%) had comorbidities. Approximately half of the respondents, 427 (50.8%), had been 2 to 4 years since the diagnosis of hypertension. About one-third of the respondents had a family history of hypertension. The majority, 730 (86.9%), had BMI measurements of 18.5 to 24.9, which were in a normal range, while only 74 (8.8%) were overweight, and the majority of the respondents, 790 (94.1%), take less than or equal to two anti-hypertensive medications (Table 2).

**Table 2:** Clinical characteristics of adult hypertension patients under follow-up at the selected public hospitals in Northwest Amhara, Ethiopia, 2022 (n=840)

| Variables |  | Frequency | Percent |
| --- | --- | --- | --- |
| Comorbidities | Yes | 243 | 28.9 |
|  | No | 597 | 71.1 |
| Time since diagnosis of HTN in years | <2 | 216 | 25.7 |
|  | 2-4 | 355 | 42.3 |
|  | ≥4 | 269 | 32 |
| Family history of HTN | Yes | 270 | 32.1 |
|  | No | 570 | 67.9 |
| BMI | Under weight | 36 | 4.3 |
|  | Normal | 730 | 86.9 |
|  | Overweight | 74 | 8.8 |
| Number of drugs taken | ≤2 | 790 | 94.1 |
|  | >2 | 50 | 5.9 |

### Individual factor of the participants

The total mean score of the 8-item modified hypertension evaluation lifestyle management (HELM) scale was used, and we used the mean score (4.62±2.11) as the cut-off point. By using this mean score as a cutoff point for more than half, 431 (51.3%) had good knowledge regarding hypertension.

### Social characteristics of the participants

Family and non-family support were evaluated using the Duke Social Support and Stress Scale. On this scale, the respondents’ overall mean score for social support was 8.5774 ± 4.03. Using this mean score as a cut-off point, about four hundred hypertensive patients (47.6%) received support from both family and non-family members, while 440 patients (52.4%) did not receive any support from either group.

### Adherence to healthy lifestyle

Regarding the overall adherence, nearly one fifth, 182 (21.7% (95% CI: 19.0–24.6)), of hypertensive patients had good adherence to diet, exercise, smoking cessation, and moderation of alcohol consumption (Fig. 2).

**Figure 2.**
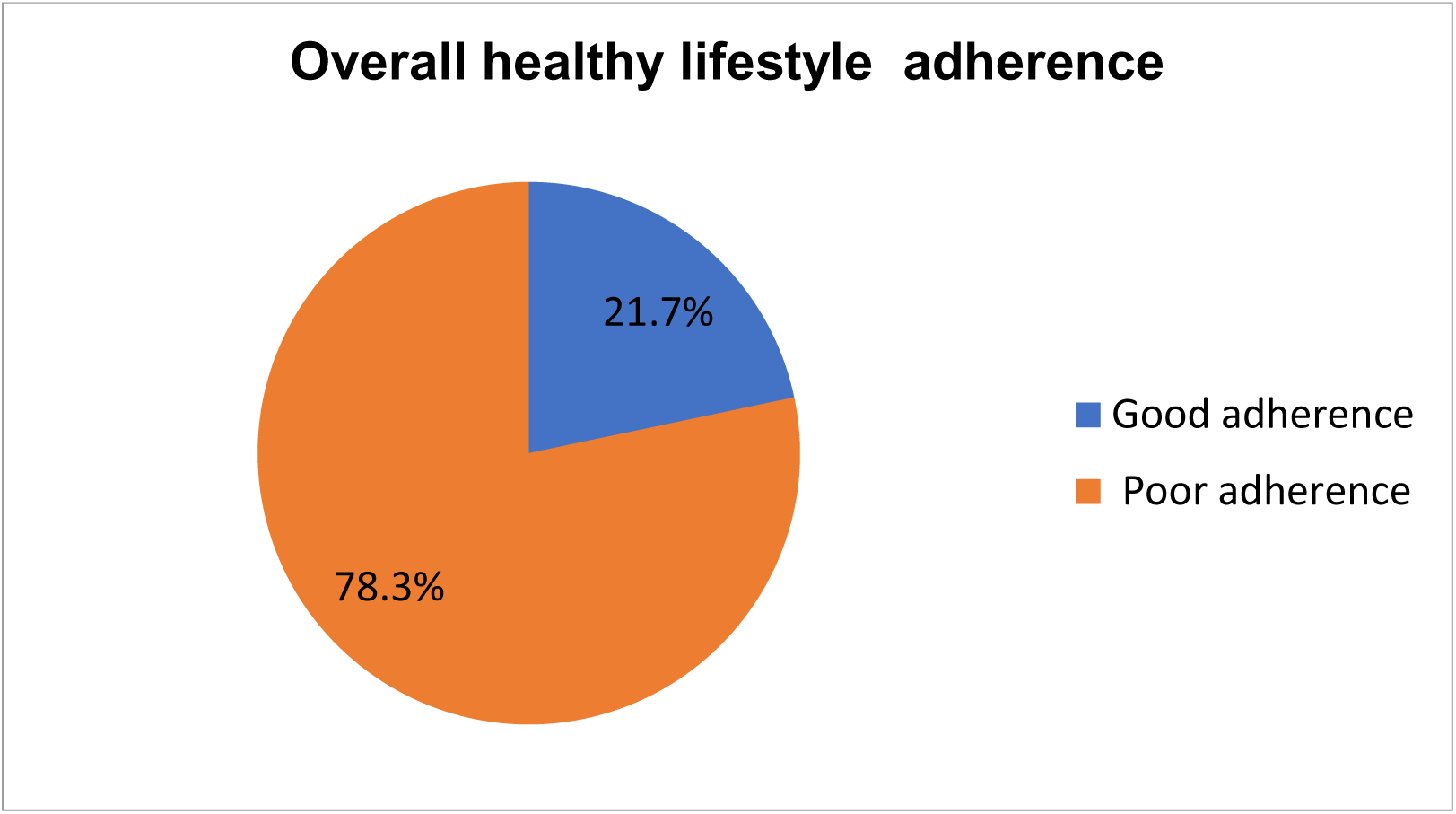
Magnitude of adherence to healthy lifestyles among adult hypertension patients under follow-up care at the public hospitals in Northwest Amhara, Ethiopia, 2022.

Of the respondents, adherence to specific domains of healthy lifestyle showed that about 388 (46.19%), 340 (40.48%), 687 (81.79%), and 555 (66.07%) were adherent to diet, physical exercise, cessation of smoking, and moderation of alcohol consumption, respectively (Fig. 3).

**Figure 3:**
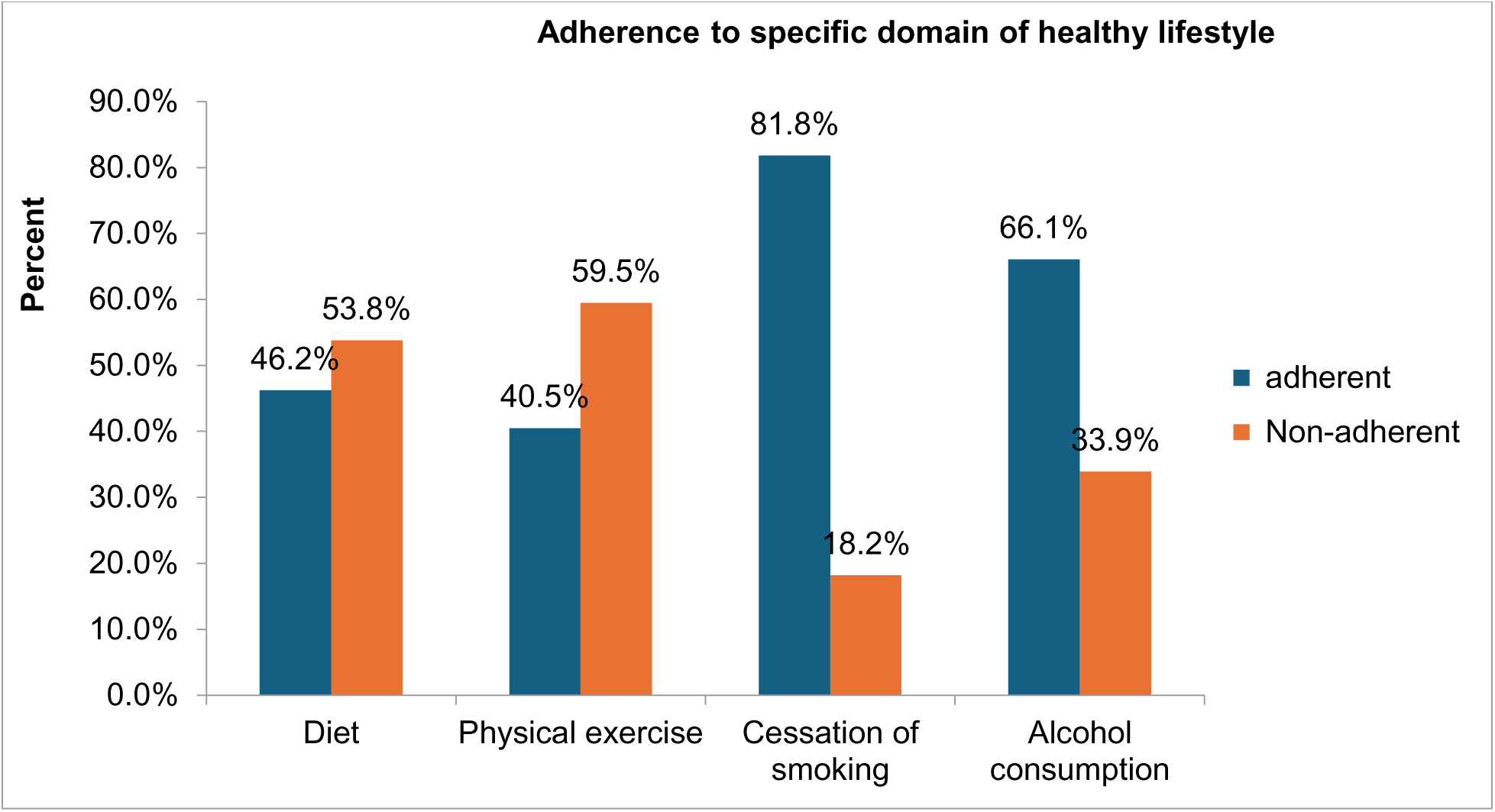
Adherence to a specific domain of healthy lifestyles among adult hypertension patients under follow-up at the selected public hospitals in Northwest Amhara, Ethiopia, 2022.

### Adherence to a diet

Since receiving a diagnosis of hypertension, 475 (56.5%) of the respondents said that they usually or always incorporated fruits, vegetables, grains, and beans in their diet, and about 88.7%, 99.1%, and 97.4% of all respondents said they had either rarely or never eaten food items high in sugar, salt, or saturated fat, respectively (Table 3).

**Table 3:** Adherence to diet among adult hypertension patients under follow-up at the selected public hospitals in Northwest Amhara, Ethiopia, 2022 (n=840)

| Variables |  | Frequency | Percent |
| --- | --- | --- | --- |
| Usually or always include fruits, vegetable, grains, and beans in their diet | Yes | 475 | 56.5 |
|  | No | 365 | 43.5 |
| **Never or rarely consume foods that contain high saturated fat** | Yes | 745 | 88.7 |
|  | No | 95 | 11.3 |
| **Never or rarely consume sugar** | Yes | 832 | 99.1 |
|  | No | 8 | 0.9 |
| **Never or rarely consume salt** | Yes | 818 | 97.4 |
|  | No | 22 | 2.6 |

### Adherence to physical exercise

Out of the total respondents, 507 (60.36%) engaged in physical activity, of whom about 354 (69.82%) exercised five days a week or more. Of the five hundred seven who performed physical exercise, walking was the dominant type of exercise performed (Table 4).

**Table 4:** Adherence to physical exercise among adult hypertension patients under follow-up at the selected public hospitals in Northwest Amhara, Ethiopia, 2022 (n=840)

| Variables |  | Frequency | Percent |
| --- | --- | --- | --- |
| **Do you perform physical exercise?** | Yes | 507 | 60.36 |
|  | No | 333 | 39.64 |
| **How often do you exercise?** | < Five days per week | 153 | 30.18 |
|  | ≥Five days per week | 354 | 69.82 |
| **For how long do you exercise per session?** | <30min | 106 | 20.91 |
|  | ≥30min | 401 | 79.09 |
| **Type of exercise do you perform?** | Walking | 330 | 65.09 |
|  | Jogging | 152 | 29.98 |
|  | Cycling | 25 | 4.93 |

### Adherence t o the cessation of smoking

The majority of respondents, 687 (81.8%), had either never smoked or had stopped. Of the 113 participants who continued to smoke cigarettes, 24 (21.2%) had tried to quit (Table 5).

**Table 5:** Adherence to cessation of smoking among adult hypertension patients under follow-up at the selected public hospitals in Northwest Amhara, Ethiopia, 2022 (n=840)

| **Variables** |  | **Frequency** | **Percent** |
| --- | --- | --- | --- |
| **Have you never smoked cigarette or stopped smoking before 30 days?** | Yes | 687 | 81.8 |
|  | No | 153 | 18.2 |
| **Have you currently smoker?** | Yes | 113 | 73.9 |
|  | No | 40 | 26.1 |
| **Have you tried to quit smoking?** | Yes | 24 | 21.2 |
|  | No | 89 | 78.8 |

### Adherence to moderation in alcohol consumption

Based on the Fast Alcohol Screening Test (FAST) score, approximately two-thirds of the respondents, 555 (66.1%), were adherent to moderation in alcohol consumption, while the remaining 285 (33.9%) engaged in harmful drinking.

### Factors associated with adherence to a healthy lifestyle

In the bi-variable logistic regression model, sex, age, residency, occupational status, educational status, monthly income, duration of hypertension diagnosis, social support, and knowledge about hypertension were found to be predictors of adherence to healthy lifestyle modification practices. After controlling for possible confounders in multivariable logistic regression, sex, educational status, duration of HTN diagnosis, knowledge about HTN, and the presence of social support were found to be statistically significant predictors of adherence to recommended lifestyle modification activities at P ≤ 0.05.

According to multivariable logistic regression analysis, being female had two times greater odds of adherence to lifestyle modification practices than being male (AOR = 2.03, 95% CI: 1.38–2.98). Hypertensive patients who attended primary education were two times more likely to adhere to recommended healthy lifestyle practices than hypertensive patients who were unable to read and write (AOR = 2.07, 95% CI: 1.06-4.03). Compared to the time of diagnosis of hypertension, hypertensive patients with 2 to 4 years of diagnosis of HTN were three times more likely to adhere to lifestyle modification practices than those with less than 2 years of diagnosis of hypertension (AOR = 3.14, 95% CI: 1.83-5.39). Participants having good knowledge about hypertension disease were three times more likely to adhere to recommended lifestyle modification practices than those who were poorly knowledgeable (AOR = 3.06, 95% CI: 2.02-4.64). Hypertensive patients getting good social support were three times more likely to adhere to recommended lifestyle modification practices than patients with poor social support (AOR = 2.89, 95% CI: 1.97–4.25) (Table 6).

**Table 6:**
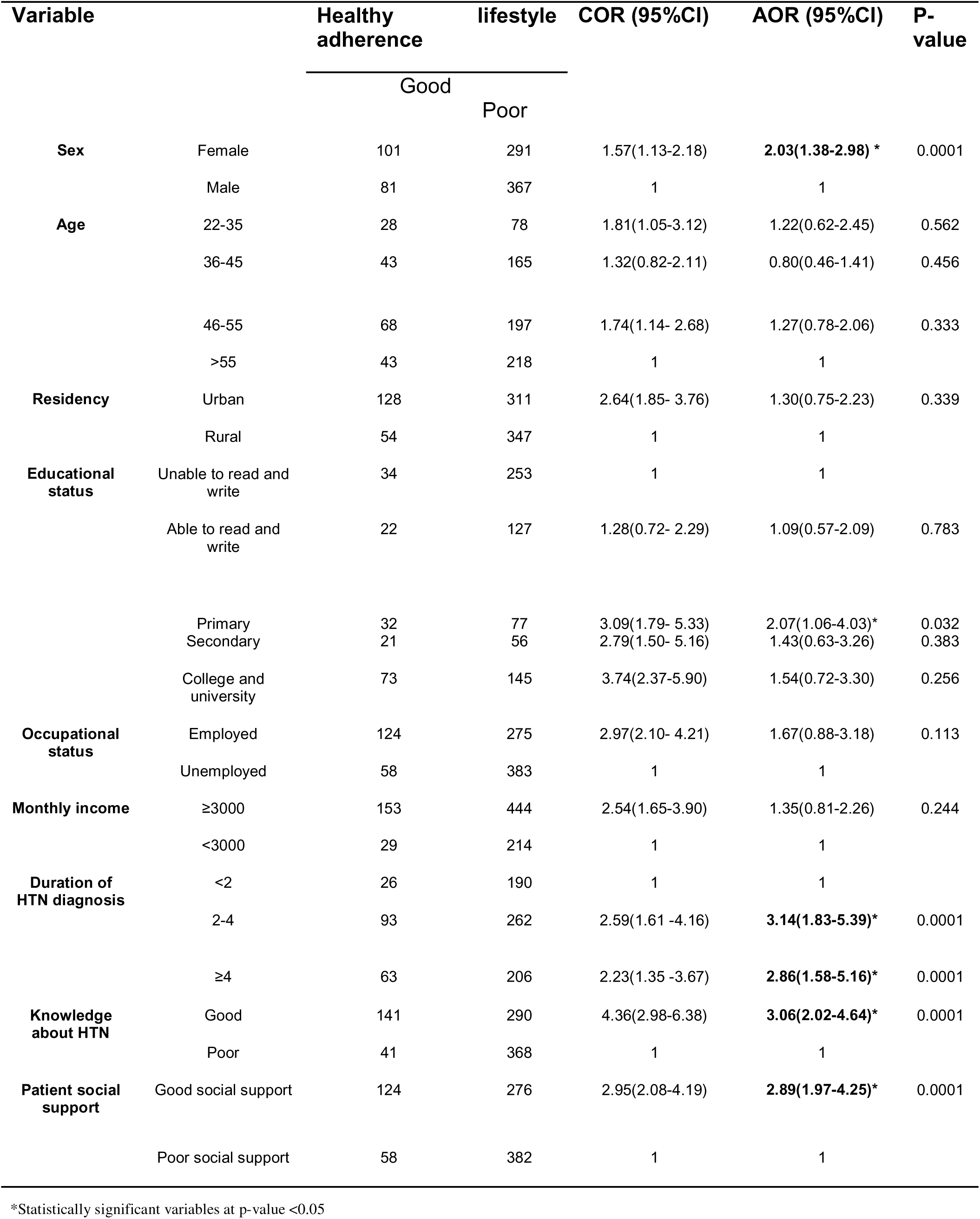
Factors associated with adherence to healthy lifestyle among adult hypertension patients under follow-up at the selected public hospitals of Northwest Amhara, Ethiopia, 2022 (n=840)

## DISCUSSION

In this study, the overall good adherence to a healthy lifestyle among hypertensive patients was 21.7% (95% CI: 19.0–24.6). This finding is in line with studies conducted in Ethiopia; Mekelle (20.3%), Dessie (23.6%), and Central Gondar (24.2%).^10,18,28^This might be due to similarities in study design and study settings, socio-demographic characteristics and lifestyles of study participants, and similar tools employed in all studies.

The current study finding is lower than studies in Ethiopia: Mizan Tepi (33.3%), Bahir Dar (32.4%), Harer (287%), Addis Ababa (46.4%), Durame and Hosanna town (27.3%), Oromia Zone, Amhara region (52.7%), Eritrea (71.8%), Ghana (72%), and India (54.7%).^3,16,17,21,29-33^ But higher than studies done in the Kingdom of Saudi Arabia (4.2%) and Nigeria (16.4%).^34-35^ The finding discrepancy might be due to differences in exposure to lifestyle modification practices, comprehensive information and knowledge, the tools used to measure lifestyle modification practices, study period, sample size difference, sociocultural and dietary habit differences, and following different lifestyle modification modalities. In India, patients were above 30 years of age, and in Saudi Arabia, only men were included in the study.

According to the multivariable regression analysis, sex, educational status, duration of hypertension since diagnosis, knowledge about hypertension, and social support were associated with adherence to a healthy lifestyle.

The likelihood of maintaining a healthy lifestyle is twice as high for females than males. Previous studies from Ethiopia, Addis Ababa, Ayder referral hospital, Tigray, and Jordan supported the current study findings.^16,26,36^ This could be because women maintain healthier lifestyles for socio-environmental and cultural reasons. For example, women may abstain from alcohol and smoking for cultural and socio-economic reasons rather than because they have hypertension.^28^ Men are more likely than women to be in settings where drinking alcohol and smoking are commonplace. Women’s lives are frequently shaped by cultural and traditional norms for modest behavior, which forbid drinking and smoking. In addition, their responsibilities as mothers and guardians of the family’s health further dissociate them from alcohol and tobacco use.^36^

Hypertensive patients who attended primary education were two times more likely to adhere to recommended healthy lifestyle practices than hypertensive patients who were unable to read and write (AOR = 2.07, 95% CI: 1.06-4.03), which is in line with a study conducted in Ethiopia, Harer, Dessie, and Central Gondar.^17,18,,28^ This might be exposure to recommended lifestyle modification practice information from various social behavioral change (SBC) tools, such as posters, leaflets, banners, billboards, radio, and television spots, and having good self-efficacy to request information from a reliable source and regularly demonstrate these lifestyles modification practices.^17^

Respondents with 2 to 4 years of diagnosis of HTN were three times more likely to adhere to lifestyle modification practices than those with less than 2 years of diagnosis of hypertension (AOR = 3.14, 95% CI: 1.83-5.39). This is in line with a study conducted in Addis Abeba.^22^ This might be due to continued counseling and health education provided by healthcare providers during follow-up visits.^29^

Those respondents who had good knowledge were three times more likely to be adherent compared to those who had poor knowledge. This is in line with a study done in Ethiopia, Addis Ababa^16^, Dessie^18^, Ayder Hospital, Tigray ^26^ and Jordan.^36^ The possible explanation could be that knowledgeable hypertension patients inquire about unclear information regarding the disease from medical professionals because they are aware of the condition, its severity, complications, and ways to prevent, control, and manage hypertension.^18^ Furthermore, referring to the health belief model and the perceived susceptibility construct theme, knowledgeable hypertension patients who received information about hypertension were more likely to follow a healthy lifestyle because they had good self-efficacy and adequately recognized the risks of hypertension.^37^

The odds of having social support from family and non-family members of society were approximately three times higher with healthy lifestyles than with poor social support. This is in line with an institutional-based study done in Ethiopia, Addis Ababa16, and Debre Tabor Hospital.^38^ Patients with social support may be more motivated to stick to healthy lifestyle modifications because these supports foster good psychological states.^39^Patients with social support from family and non-family members demonstrated good adherence than with no social support, which is supported by health belief model construct of enhanced self-efficacy where encouragement from family strengthens confidence in overcoming barriers to lifestyle change.^37^

## STRENGTH AND LIMITATION OF THE STUDY

Levels of social support were assessed by using eleven items on the Likert scale and the Duke Social Support and Stress scale. In addition, the Fast Alcohol Screening Test (FAST), which is the short version of the alcohol disorders identification test (AUDIT), was also used to assess adherence to alcohol consumption, which was inherited to recall bias. However, these scale measurements are a widely used instrument in cross-sectional studies to measure alcohol use and social support levels. Despite these limitations, it is a preliminary study in the study areas that contributes to the scant body of evidence by offering helpful information for hospitals, medical professionals, and hypertension patients.

Other drawbacks of the study were that the diet measuring tool was subjective to quantify exactly what participants consumed during the data collection period, and the questionnaire employed in this study might be prone to social desirability bias. Moreover, other limitations of this study include the cross-sectional nature of the study design, which makes it difficult to establish a causal relationship between following a healthy lifestyle recommendation and related characteristics.

## CONCLUSION

The overall adherence to recommended healthy lifestyle modification practices among hypertension patients in this study was low. Independent predictors of adherence to lifestyle modification practices were sex, educational status, duration of hypertension diagnosis, knowledge about hypertension, and social support. Therefore, the Amhara region health bureau and other non-governmental organizations working on chronic illness including hospital manager should arrange capacity-strengthening training on recommended healthy lifestyle practices for health workforce. Health care professionals should provide special attention and proper and adequate counseling for hypertensive patients with low adherence to recommended healthy lifestyle modifications and low knowledge about hypertension. Furthermore, the facility should assign a full or part-time social worker for proper counseling and support for patients with poor social support.

## List of abbreviations

AOR: adjusted odds ratio
BMI: Body Mass Index
BP: Blood Pressure
CI: Confidence Interval
COR: Crude Odds Ratio
CVD: Cardio-Vascular Diseases
CSA: Central Statistical Agency
FAST: Fast Alcohol Screening Test
HELM: Hypertension Evaluation of Lifestyle and Management
HTN: Hypertension
NCD: Non-Communicable Diseases
SBC: Social Behavioral Change
VIF: Variance Inflation Factor
WHO: World Health Organization

## Acknowledgement

We express our gratitude to the University of Gondar for granting us the ethical clearance needed to carry out this study. We extend our sincere gratitude to the patients who took part in the study, supervisors, hospital managers, and data collectors.

## Authors Contributions

MZ, GAB, DZ and AAM contributed to the conception, design of the study, and supervision during data collection. MZ, GAB, MG, HGZ, and AW worked on the formal analysis and methodology and drafted the manuscript. AAM, GAB, TA, TED and HGZ critically reviewed the draft manuscript, wrote the final version and participated to address feedbacks. All authors read and approved the final manuscript.

## Patient consent for publication

Consent obtained directly from patient(s).

## Ethics approval and informed consent

The University of Gondar’s Institute Review Board (IRB) granted ethical approval. A support letter was obtained from the University of Gondar, College of Medicine and Health Science, Institute of Public Health, to be given to the respective hospitals. Confidentiality of the participants was ensured by excluding the participants’ names and other identifiers in the data abstraction format. The participants were informed that they had the right to withdraw from the study at any point in the interview. After the purpose of the study was explained, written informed consent was obtained from each study participant.

## Data availability statement

All data and materials are available from the corresponding author without undue restriction. All data relevant to the study are included in the article or uploaded as supplementary information.

